# Quantitative sodium-MRI detects differential sodium content in benign vs. malignant oncocytic renal tumours

**DOI:** 10.1101/2024.06.19.24309026

**Authors:** Ines Horvat-Menih, Jonathan R Birchall, Maria J Zamora-Morales, Alice Bebb, Joshua D Kaggie, Frank Riemer, Andrew B Gill, Andrew N Priest, Marta Wylot, Iosif A Mendichovszky, Anne Y Warren, James Jones, James N Armitage, Thomas J Mitchell, Grant D Stewart, Mary A McLean, Ferdia A Gallagher

## Abstract

**Background:** Accurate non-invasive subtyping of localised kidney tumours is an unmet clinical question in uro-oncology. Differentiation of benign renal oncocytomas (RO) from malignant chromophobe renal cell carcinomas (chRCC) is not possible using conventional imaging. Despite the importance of renal function for sodium regulation, little is known about sodium handling in kidney tumours.

**Purpose:** Here we used non-invasive sodium MRI (^23^Na-MRI) to quantify sodium concentration and relaxation dynamics across a range of different kidney tumour subtypes and have correlated these findings with imaging surrogates for perfusion, hypoxia, and cellularity.

**Materials and Methods:** Between January and April 2023, patients with localised renal masses were prospectively recruited and underwent ^23^Na and proton (^1^H) MRI at 3T to acquire 3D maps of B_1_, total sodium concentration (TSC), proton and sodium relaxation rates (*R*_2_*), and diffusion weighted imaging (DWI). Statistical analysis included comparison and correlation of quantified imaging parameters across kidney tumour subtypes.

**Results:** Ten patients were included in the final analysis (mean age±S.D. = 64±8 years; 7:3 male:female ratio) encompassing seven ROs, two chRCCs, two clear cell RCCs (ccRCC), and one papillary RCC (pRCC). The TSC was significantly higher in the ROs compared to the chRCCs: 162±58 mM vs. 71±2 mM (*P* < 0.05). The mean TSC in ccRCC was 135±59 mM, and 81 mM in pRCC. The ^23^Na-derived and ^1^H-derived *R*_2_* values showed a weak correlation (Spearman r = 0.17; *P* = 0.50). There was a significant inverse correlation between TSC and ^1^H-*R*_2_* (Spearman r = -0.39, *P* < 0.05), but TSC was independent of the DWI-derived imaging parameters.

**Conclusion:** ^23^Na-MRI detected markedly different sodium concentrations within benign ROs and malignant chRCCs. In addition, the sodium signal inversely correlated with ^1^H-*R*_2_* as a surrogate for hypoxia. Therefore we have shown the feasibility and potential of ^23^Na-MRI for future research in renal tumours.

**Key results:**

1. ^23^Na-MRI was used to non-invasively assess kidney tumour subtypes for the first time.
2. A significantly higher total sodium concentration was detected in benign renal oncocytoma (162±58 mM), compared to chromophobe renal cell carcinoma (71±2 mM), as the malignant counterpart: *P* < 0.05.
3. Total sodium concentration showed a significant inverse correlation with ^1^H-*R*_2_* (Spearman r = -0.39, *P* < 0.05), but it was independent of the diffusion-weighted imaging-derived parameters.

**Summary statement:** ^23^Na-MRI showed potential for differentiating benign and malignant masses, to characterise kidney tumours, which may be linked to the underlying differences in deoxygenation as measured with ^1^H-MRI.

## Introduction

Kidney tumours pose an increasing health economic burden due to the rising incidental detection of small renal masses (SRMs), which are typically ≤ 4cm^1^. These SRMs pose a diagnostic dilemma as they often remain indeterminate on initial evaluation due to low specificity of current clinical imaging methods^2^. This diagnostic uncertainty may result in unnecessary surgery, with the potential loss in renal function for those patients undergoing inappropriate nephrectomy. Conversely, a long-term active surveillance strategy with imaging may be more appropriate than surgical resection for many patients, but this can affect patient mortality and morbidity if the detection of a malignant tumour is delayed, and equally can result in psychological morbidity for patients with benign disease as they await a diagnosis^3^. Renal mass biopsy prior to the decision for surgery can aid in the identification of benign cases, but misdiagnosis may result due to the challenges of accurately sampling small lesions, particularly in the context of tumour heterogeneity^4^. Novel non-invasive imaging techniques could aid the characterisation of renal masses on a whole-tumour level.

Functional MRI methods enable tumour biology to be probed beyond simple morphological evaluation and therefore aid differentiation and stratification, as exemplified by the recently developed clear cell likelihood scoring system (ccLS)^5^. While the ccLS requires qualitative interpretation by a radiologist, quantitative MRI biomarkers have the potential of providing objective, robust, quantitative comparisons between patients^6^. Despite the potential of these functional MRI metrics for clinical benefit, there have been few published studies in this field which mostly report surrogates for perfusion^7^, cellularity^8^, and hypoxia^9^, and all require further investigation.

Regulation of plasma and urinary sodium concentration is a primary renal function, but the role of sodium and its concentration gradient within kidney tumours is less well understood^10,11^. Many cancers exhibit alterations in ion dynamics to maintain an alkaline intracellular pH and acidic extracellular pH, which promotes survival and tumour aggressiveness^12^. Furthermore, it has been suggested that activation of the sodium hydrogen exporter (NHE1) can be the initial step driving development of other cancer hallmarks such as the Warburg effect and increased cell proliferation^13^. Therefore, detecting the sodium phenotype of tumours may provide an insight into initial genetic drivers and *in vivo* hypoxic and pH-influences, which may aid the characterisation of kidney tumours.

Sodium MRI (^23^Na-MRI) enables the non-invasive quantification of spatial and temporal alterations in sodium concentrations^14,15^. In oncological imaging, ^23^Na-MRI has been shown to detect higher sodium concentrations in tumours compared to the normal-adjacent prostate tissue^16^, and in ovarian cancer the total sodium concentration correlated negatively with cell density^17^. In human kidneys, ^23^Na-MRI could be used to detect the corticomedullary sodium gradient and its responsiveness to diuretics^18^, as well as the differences between healthy and diseased kidneys^19^. Here we set out to explore the role of ^23^Na-MRI in renal mass characterisation and to assess sodium concentration in benign and malignant renal masses. The underlying biological changes driving this were assessed by correlating the ^23^Na-MRI signal with factors influencing sodium regulation *in vivo*, including deoxygenation, perfusion, and diffusion as probed with multiparametric MRI.

## Patients and Methods

### Ethics and patient recruitment

Consecutive patients with localised renal masses were prospectively recruited in the uro-oncology clinic in Addenbrooke’s Hospital, Cambridge University Hospitals NHS Foundation Trust, Cambridge, UK between January and April 2023, and signed the informed consent for the feasibility study approved by the institutional review board (Research Ethics Committee number: 22/EE/0136). The main inclusion criteria were: ≥18 years of age, clinical suspicion of renal mass, and an Eastern Cooperative Oncology Group (ECOG) performance status ≤1. Key exclusion criteria were unsuitability for MRI, significant comorbidities, pregnancy, and histological subtype other than those originating from renal tubular epithelium. Histopathology of the renal mass was determined either on biopsy or on the surgical specimen, depending on the clinical decision.

### ^23^Na-MRI acquisition and analysis

Recruited patients underwent ^23^Na-MRI on a clinical 3T scanner (Discovery MR750, GE Healthcare, Waukesha, WI), using a large field of view (FOV) ∼48 cm ^23^Na-tuned transmit/receive birdcage coil (Rapid Biomedical GmbH, Rimpar, Germany)^20^. High-resolution sodium images were acquired with a 3D cones trajectory^21^, using the following acquisition parameters: echo time (TE) 0.705 ms, repetition time (TR) 150 ms, flip angle 70°, voxel size 4 × 4 × 8 mm, 1402 transients, 5 averages, receiver bandwidth 167 kHz, and 11 min 41 s duration. A pair of 80 mM agar gel NaCl phantoms placed within the imaging FOV served as a calibration standard for signal intensity^22^. B_1_ maps were acquired using the double angle method (DAM) as previously described^23^. For patients with phantoms placed close to the edges of the FOV and thus unreliable B_1_-correction, a population average estimate of local B_1_ intensity based on healthy volunteer data was used as a substitute. Low resolution 3D sodium cones images (TR 100 ms, flip angle 70°, voxel size 9 x 9 x 9 mm, 197 transients, 4 averages, receiver bandwidth 125 kHz, 1 min 19 s duration per series) were also acquired at six different echo times (TEs = 0.7, 1.5, 2, 4, 8, 16 ms) to estimate the sodium *T*_2_* values. Anatomical fat/water proton MRI was acquired with a 3D two-point Dixon sequence using the MR system-integrated body coil, within a single breath hold: TEs 1.1/2.2 ms, TR 3.7 ms, flip angle 15°, 1 average, matrix size 256 x 192, 0.7 phase FOV, 40 cm FOV.

Maps of the uncorrected total sodium concentration (TSC) were generated relative to an 80 mM agar gel ^23^Na-phantom placed within the same FOV using MATLAB (MathWorks, Natick MA). Maps of monoexponential estimates of the ^23^Na *T*_2_* relaxation time were generated from linear fitting of log-transformed intensities of ^23^Na signal in the variable-TE series. ROIs within tumours, normal-appearing kidney parenchyma and liver were drawn on 2D axial slices in each parameter map by a resident in-training with 4 years of experience in kidney segmentation, supervised by a consultant academic radiologist with more than 20 years of experience. Mean signal intensities within these ROIs were extracted. As an alternative method for estimation of TSC - to be applied in situations where the phantom signal was considered unreliable for use as a reference - further normalisation was performed using the ^23^Na signal from the liver as a reference, that is by dividing the uncorrected TSC from each ROI with the TSC from the liver region within each patient. Transmit and receive B_1_ uniformity correction was performed on a per-region basis to obtain estimates of B_1_-corrected TSC per region.

### ^1^H-MRI acquisition and analysis

After the sodium acquisition with the birdcage transmit/receive coil, a change of coil (32-channel cardiac array coil, GE Healthcare, Waukesha, WI) was necessary to perform proton *R*_2_* mapping and IVIM DWI. *R*_2_* mapping was undertaken using the images acquired with the following parameters: FOV 40 cm, 12 echo times with 3.1 ms spacing; TR 110 ms, flip angle 30°, matrix size 256 × 224, slice thickness 4 mm, multiple breath holds, echo train length 12; and processed as previously described^24^. IVIM DWI was acquired using a respiratory-triggered dual spin-echo echo-planar imaging sequence and b-values of 0, 10, 20, 30, 100, 300, 500, 700 and 900 s/mm^2^ with the following parameters: TE ∼80 ms, TR 1 respiratory cycle, FOV 28.8 cm, slice thickness 4 mm, acquisition matrix 96 × 96, 2 averages for b-values <100 s mm^-2^ and 4 averages for higher b-values, receiver bandwidth ±111 kHz, parallel imaging (ASSET: Array coil Spatial Sensitivity Encoding) factor 2, averaged 3 directions, acquisition time 91 breaths (∼10 min). MATLAB was used for processing of the IVIM DWI data to extract the perfusion fraction (*f*_p_) and the diffusion coefficient (*D*_0_), as previously described^24^.

OsiriX MD v.14.0 (Pixmeo Sarl, Bernex, Switzerland) software was used to reorient and register the proton maps in the same space as the ^23^Na-MRI acquisition, and quantification of the mean values for individual parameters was performed using the same ROIs as annotated for ^23^Na-MRI analysis.

### Statistical analysis

Statistical analysis was performed in GraphPad Prism v.10. Normality of the data was tested using the Shapiro-Wilk test. Normally distributed data and results are presented as mean (±S.D., standard deviation), and non-parametric data were presented as median (range). Welch’s unpaired t test and Welch’s ANOVA test were used for comparison of normally distributed imaging parameters across the kidney tumour subtypes, and for nonparametric parameters, the Kruskal-Wallis with Dunn’s multiple comparisons test was used. *P* < 0.05 was used as the cut-off for significance. Sample size was determined based on power calculation as per IRB-approved protocol.

## Results

### Patient characteristics

A summary of the patient characteristics is shown in Table 1. Seven renal oncocytomas (RO), two chromophobe RCCs (chRCC), two clear cell RCCs (ccRCC), and one papillary RCC (pRCC) were identified across the 10 participants with mean age±S.D = 64±8 years, of which 7 were men. Figure 1 presents the flow diagram of excluded participants: from the initial twelve recruited participants, two were excluded from the final analysis due to unsuitable histology: one bearing an angiomyolipoma (AML), and one patient bearing a tumour which remained indeterminate due to the decision for active surveillance without gaining histology with a renal tumour biopsy.

**Table 1:** Patient characteristics.

|  |  |
| --- | --- |
| **Age at detection**<br>(mean±S.D.) [years] | 64±8 |
| **Sex**<br>(Female:male) | 3:7 |
| **Presentation at detection**<br>(Incidental / symptom-triggered investigation) | 10/0 |
| **Tumour size at detection**<br>(mean, S.D.) [cm] | 4.4±1.6 |
| **Histology** |  |
| RO | 6 (including one bilateral) |
| chRCC | 2 |
| ccRCC | 3 |
| pRCC | 1 |
| **Clinical management** |  |
| Active surveillance | 5 (including one bilateral) |
| Nephrectomy | 6 |

S.D. = standard deviation, RO = renal oncocytoma, RCC = renal cell carcinoma, chRCC = chromophobe RCC, ccRCC = clear cell RCC, pRCC = papillary RCC.

**Figure 1:**
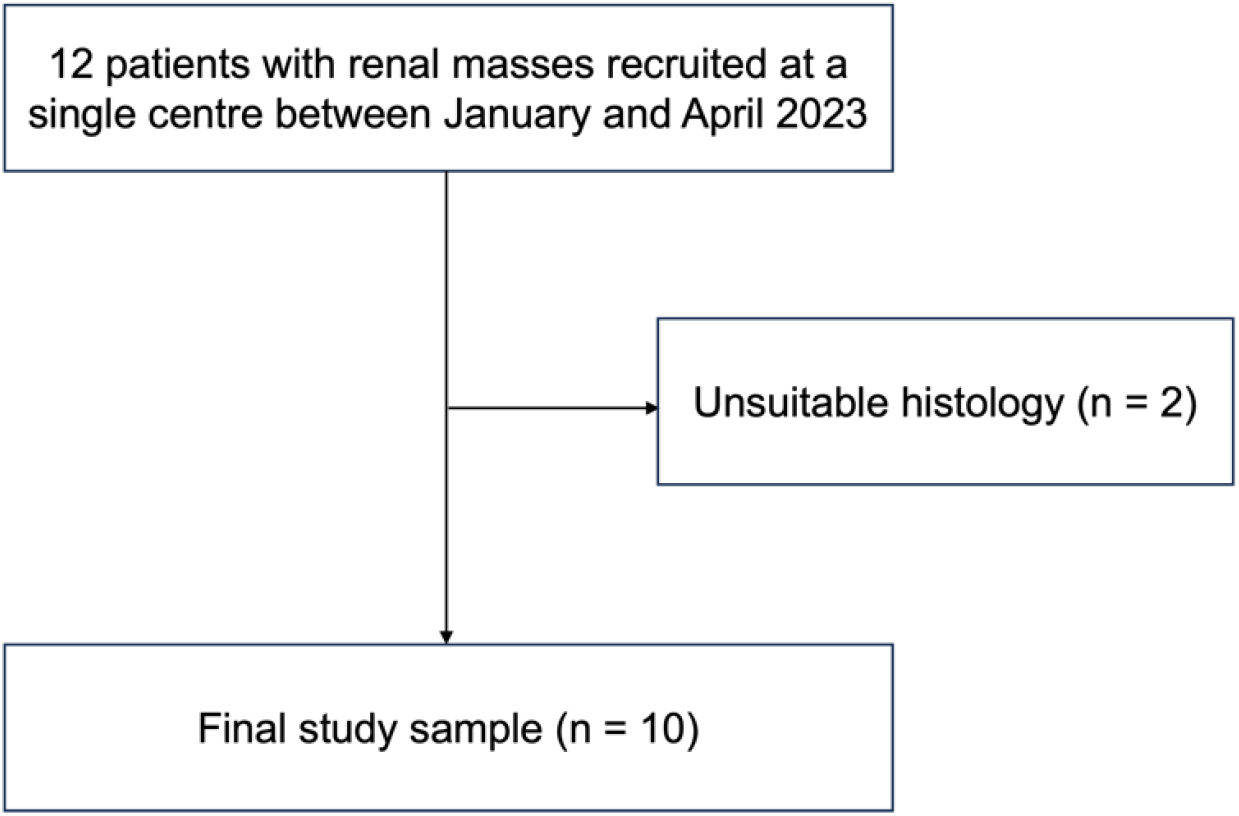
Flowchart of patient inclusion.

Initial detection of the renal mass was incidental in all participants i.e. imaging was undertaken for reasons other than renal pathology. The mean tumour size at detection was 4.4±1.6 cm. Four patients were on active surveillance at the time of their research MRI and remain on active surveillance; therefore, post-operative tissue analysis is unavailable. Six patients underwent nephrectomy shortly after their research imaging, with one patient undergoing nephrectomy for a right-sided ccRCC whilst retaining a left-sided chRCC which continues to be followed with active surveillance.

### ^23^Na-MRI: TSC and R_2_* quantification across kidney tumour subtypes

Representative maps of sodium signal intensity overlaid on T_1_-weighted anatomical images in examples of the evaluated kidney tumour subtypes are shown in Figure 2. B_1_-corrected, phantom-normalised TSC results are summarised across kidney tumour subtypes in Figure 3a, and liver-normalised TSC results are presented in Supplementary Figure 1. TSC quantifications per patient are summarised in Supplementary Table 1.

**Figure 2:**
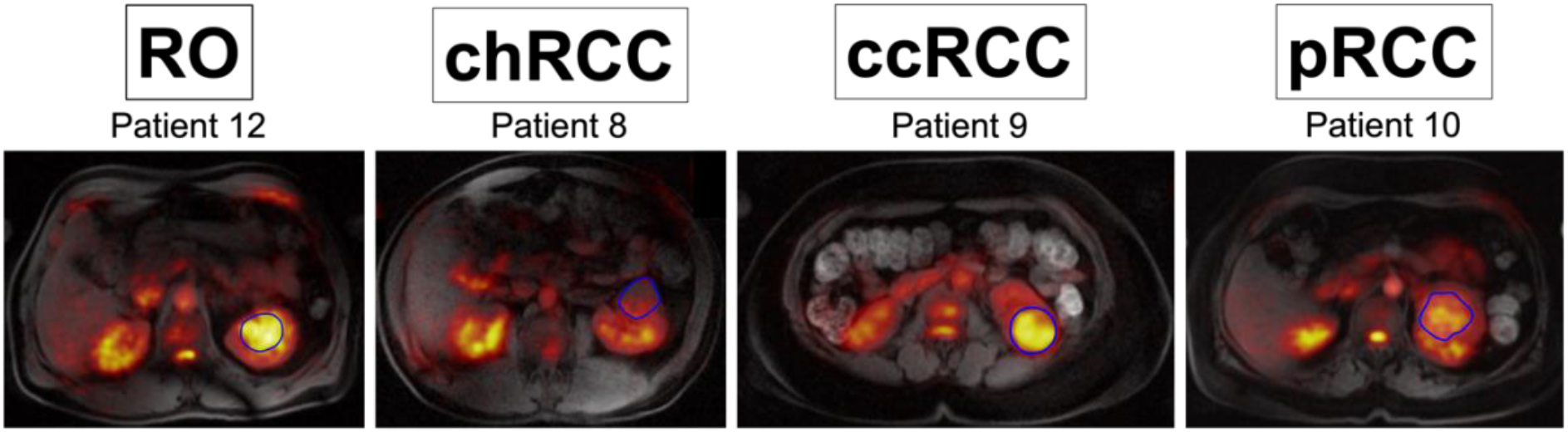
Representative maps of sodium signal intensity overlaid on T_1_w images for the evaluated kidney tumour subtypes annotated in blue. RO = renal oncocytoma, RCC = renal cell carcinoma, chRCC = chromophobe RCC, ccRCC = clear cell RCC, pRCC = papillary RCC.

**Figure 3:**
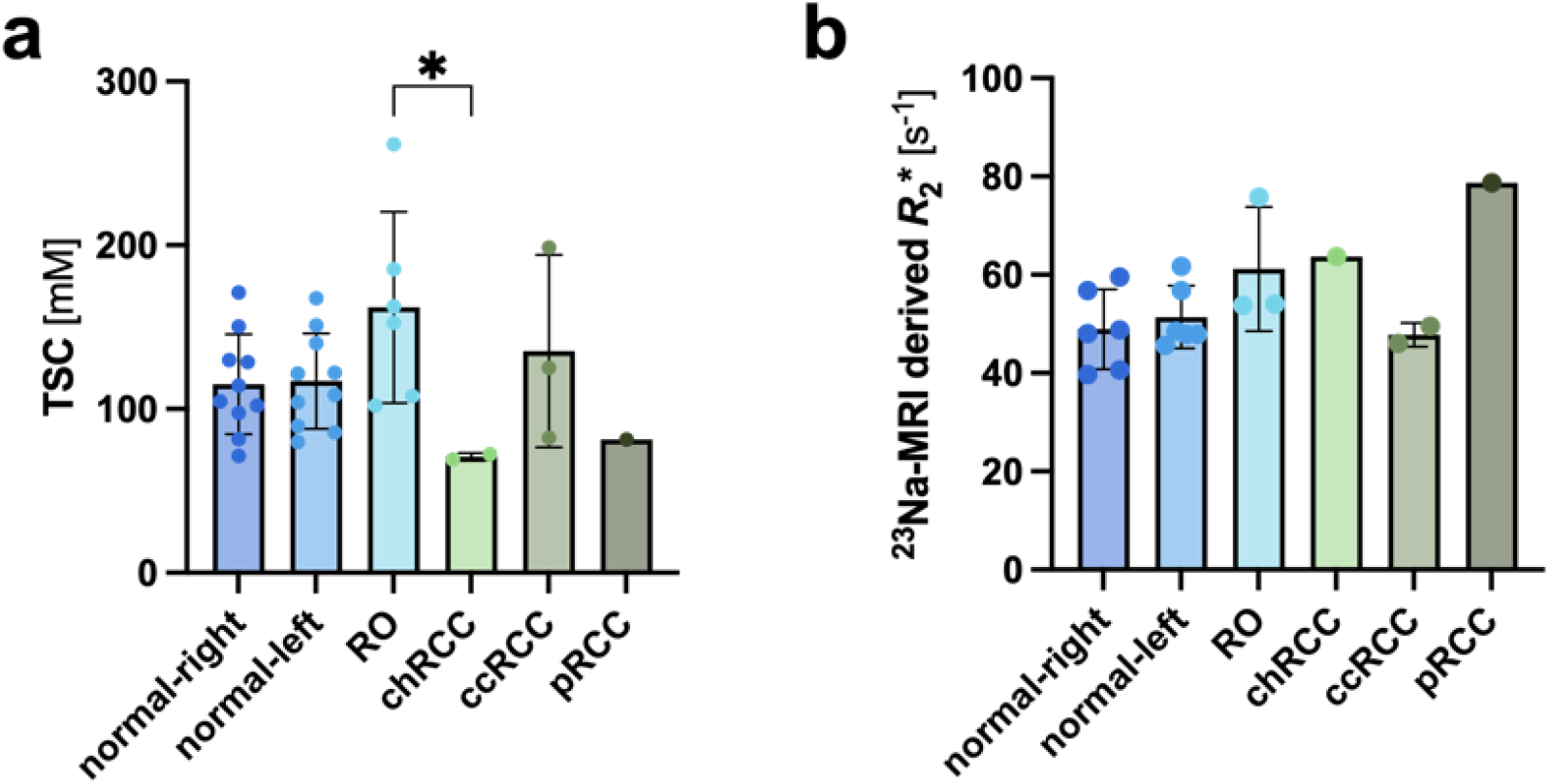
Barplots representing the B_1_-corrected mean (a) TSC and (b) *R*_2_* in normal kidney parenchyma and across the kidney tumour subtypes. Welch’s ANOVA test presented as * *P* < 0.05.

The mean TSC of the normal kidney parenchyma was similar bilaterally (right: 115±31 mM; left: 117±29 mM) with the highest TSC observed in the benign RO (162±58 mM), followed by the ccRCC (135±59 mM) and the lowest TSC measured in chRCC (71±2 mM), followed by the pRCC (81 mM). The comparison between RO and chRCC was found to be statistically significant (*P* < 0.05), and the significance remained even when considering the outlier in RO dataset (RO: 142±36 mM vs. chRCC: 71±2 mM; *P* < 0.05). TSC quantifications normalised to liver showed similar trends to those observed using phantom normalisation; however, the differences missed statistical significance by a small margin (Kruskal-Wallis with Dunn’s multiple comparisons correction, *P* = 0.08).

*R*_2_* mapping of the ^23^Na-nucleus was performed in 6 patients; Figure 3b shows the comparison of B_1_-corrected *R*_2_* across kidney tumour subtypes, with the full results in Supplementary Table 2. Interestingly, the trend in ^23^Na-MRI derived *R*_2_* showed a different trend compared to the TSC results, with highest values detected in the pRCC (79 s^-1^), followed by chRCC (64 s^-1^), RO (61±13 s^-1^) and ccRCC (48±2 s^-1^). *R*_2_* values obtained from normal kidney parenchyma were similar bilaterally (right: 49±8 s^-1^, left: 51±6 s^-1^). ANOVA analysis was not applied due to single data points for chRCC and pRCC, while no statistical significance was observed in Welch’s t test comparing RO and ccRCC (*P* = 0.20).

### Comparison of R_2_* values derived from ^23^Na-MRI and ^1^H-MRI

^23^Na-MRI derived *R*_*2*_*\** values were compared to those derived from ^1^H-MRI, with representative images from different kidney tumour subtypes shown in Figure 4. Supplementary Table 2 summarises the quantitative results from the patients who underwent *R*_2_* mapping. The absolute values were of different magnitudes (range between 40-79 s^-1^ for ^23^Na-MRI derived *R*_*2*_*\** (Figure 3b), and between 12-48 s^-1^ for ^1^H-MRI derived *R*_*2*_*\** (Figure 5a)).

**Figure 4:**
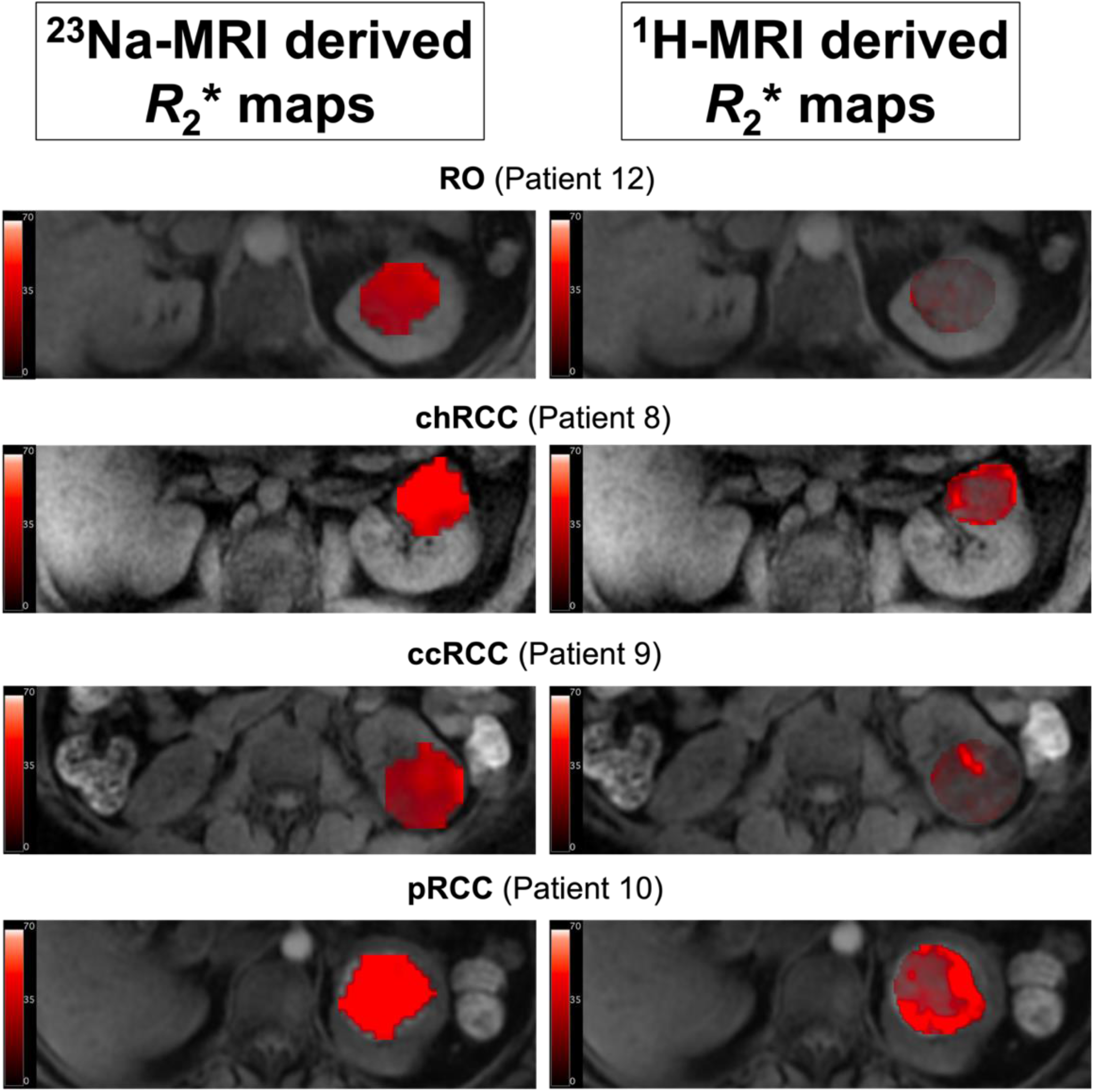
Representative *R*_2_* maps derived from ^23^Na-MRI (left side) and ^1^H-MRI (right side), with tumour overlays on T_1_w anatomical image. Units of the scale bar = s^-1^.

**Figure 5:**
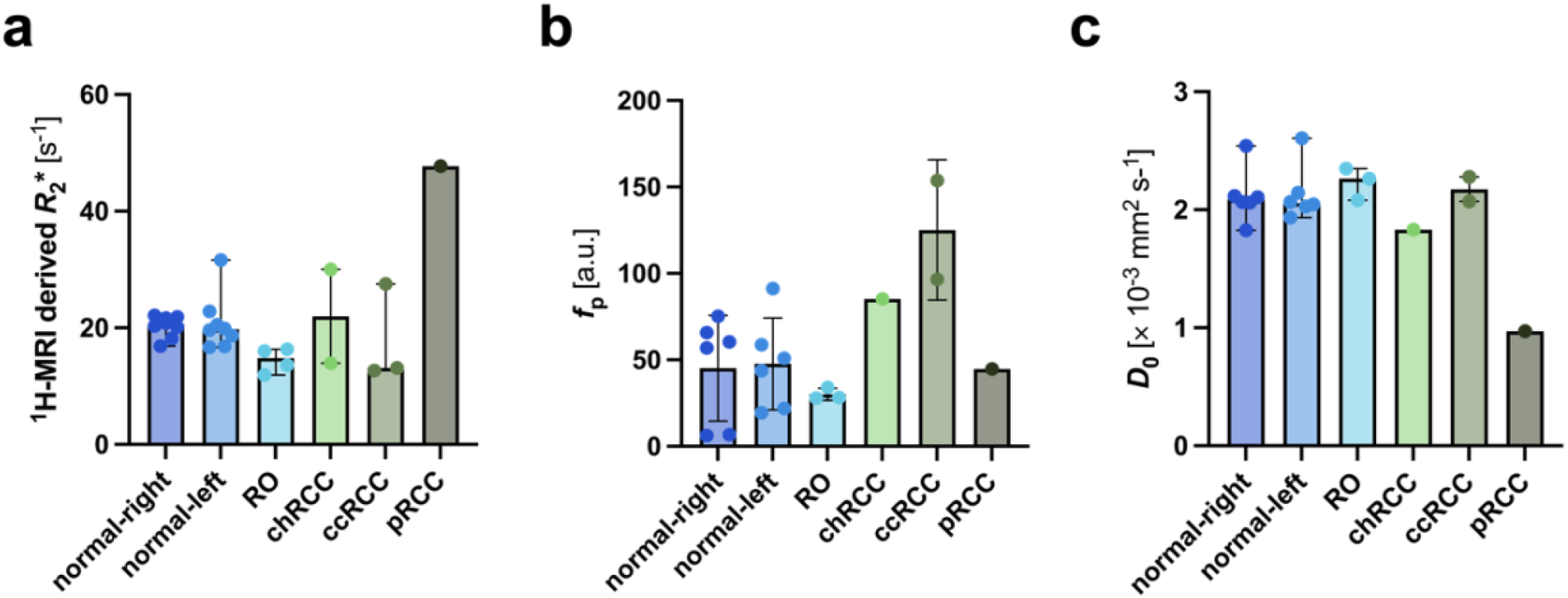
Barplots depicting ^1^H-MRI results including (a) *R*_2_* [median (range)], (b) *f*_p_ [mean±S.D.] and (c) *D*_0_ [median (range)] across kidney tumour subtypes and normal parenchyma.

The highest observed *R*_*2*_*\** values were observed in the single pRCC tumour subtype from both ^1^H- and ^23^Na-MRI. Similarly, the ccRCC tumour subtype exhibited the lowest *R*_*2*_*\** values using both techniques.

The largest difference between the *R*_*2*_*\** values derived from the two nuclei was identified in the RO group: ^23^Na-MRI derived *R*_*2*_*\** in this tumour type was among the highest (61±13 s^-1^), while *R*_2_* derived from ^1^H-MRI was among the lowest (15, range 12-16 s^-1^). Kruskal-Wallis with Dunn’s multiple comparisons test applied to the ^1^H-MRI-derived *R*_*2*_*\** data across tumour subtypes was not statistically significant (*P* = 0.34).

### IVIM DWI results

IVIM DWI was acquired in the same 6 patients as the *R*_2_* mapping. Figure 5 represents the quantitative results for *f*_p_ (perfusion fraction) and *D*_0_ (diffusion coefficient), comparing different kidney tumour subtypes, with detailed results found in Supplementary Table 3.

The lowest perfusion measurements (*f*_p_) and the same time highest diffusion surrogate (*D*_0_) was recorded in the RO group: 0.031 (0.004) and 2.264 (2.083-2.348) × 10^−3^ mm^2^ s^-1^, respectively. The ccRCC showed the highest *f*_p_ (0.125±0.041), while pRCC exhibited the lowest *D*_0_ (0.970 × 10^−3^ mm^2^ s^-1^). However, none of the comparisons was deemed to be statistically significant (*f*_p_: Welch’s t test, *P* =0.18; *D*_0_: Kruskal-Wallis with Dunn’s multiple comparisons test, *P* = 0.30).

### Correlations between imaging parameters

To understand the influence of the imaging-derived indicators of tumour macroenvironment on the TSC, a correlative analysis was performed with the correlation matrix shown in Figure 6.

**Figure 6:**
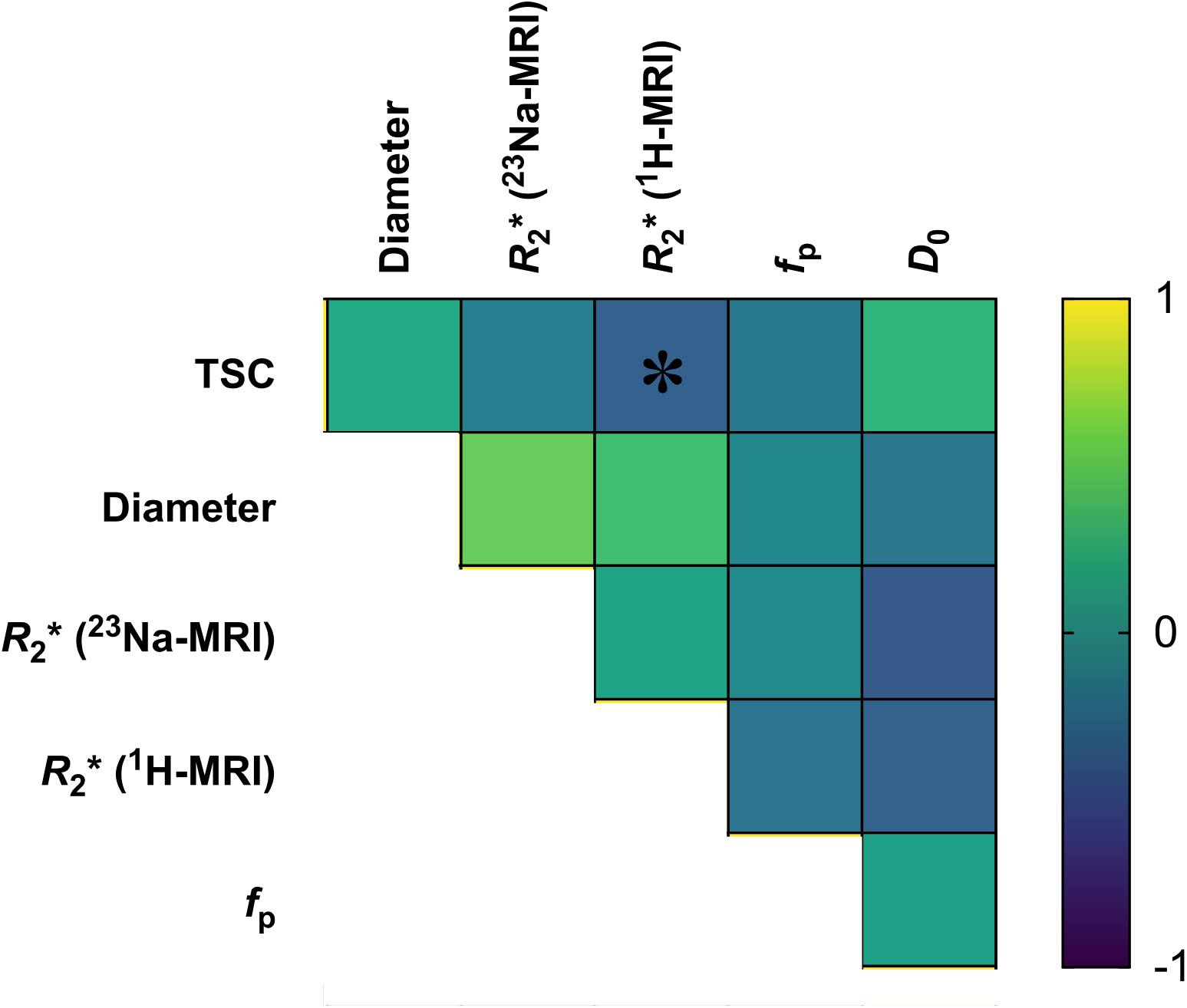
Correlation matrix of imaging parameters. Spearman correlation significance presented as * *P* < 0.05.

TSC showed a significant negative correlation with the ^1^H-derived *R*_2_* (Spearman r = -0.39, *P* < 0.05), but not with ^23^Na-derived *R*_2_*. The correlation between both *R*_2_* parameters was not found to be statistically significant (Spearman r = 0.17, *P* = 0.50), and no other correlations fell within significance.

## Discussion

Accurate non-invasive subtyping of small renal tumours is a major unmet clinical challenge in uro-oncology, which has significant implications for patient mortality and quality of life, as well as the economic burden on healthcare systems^25^. Developing novel imaging methods to characterise and stratify indeterminate kidney tumours at an early stage with high sensitivity and specificity has important potential^3^. In this study we investigated the role of ^23^Na-MRI in kidney tumour subtyping showing the potential of methods such as B_1_-correction and *R*_2_* mapping, as well as comparing to other biological imaging biomarkers for perfusion, cellularity, and hypoxia.

This is the first report of measuring the sodium concentration in kidney tumours using ^23^Na-MRI, with previous studies focusing on the role of the technique in diffuse kidney disease^26^. The quantified TSC values within normal kidney parenchyma were within a range that is similar to those reported previously^18,27^. Most notably, we detected a significantly higher TSC in the benign ROs compared to the chRCCs as its malignant counterpart: differentiating these two tumour types is a major clinical challenge due to their similarity on conventional imaging as well as on biopsy, and emerging research imaging methods are unable to distinguish between the two^28^. The higher TSC in RO compared to chRCC may be partly explained by lower cellularity and an increased extracellular volume fraction, measured as a higher *D*_0_ in the former, although this was not statistically significant. We have previously observed in ovarian cancer a similar inverse correlation between TSC and cellularity measured on histology^17^.

We detected an inverse relationship between the TSC and *R*_2_* as measured with ^1^H-MRI, but not with the ^23^Na-MRI derived *R*_2_*, suggesting that the different multinuclear *R*_2_* measurements are probing different aspects of biology. We hypothesise this to be due to differences in the compartmentalisation of sodium within the tissue compared to free water as well as the quadrupolar relaxation properties of sodium, compared to protons^29^. Due to the sample size, we were not able to assess the correlation between TSC with *D*_0_ in each of the tumour subtypes individually, although a previous study in healthy human kidney found that the TSC correlated with neither apparent diffusion coefficient (ADC) nor with *R*_2_*^30^.

The ^23^Na-*R*_2_* and ^1^H-*R*_2_* values that we reported within the normal kidneys agreed with existing literature^30,31^, but only ^1^H-*R*_2_* measurements have been reported in kidney tumours^9^. Wu et al. (2015)^9^ observed a higher ^1^H-*R*_2_* in benign lesions compared to RCC, but did not report results for individual kidney tumour subtypes, such as both benign lesions (RO and AML), nor the three main RCC subtypes. Choi et al. (2014)^32^ observed the highest ^1^H-*R*_2_* in chRCCs across the three RCC subtypes, which was in agreement with our ^1^H-*R*_2_* measurements, but they did not compare this to benign lesions.

*R*_2_* on ^1^H-MRI is often used as a biomarker or surrogate of hypoxia^33^, but is also affected by other changes in tissue such as iron overload^34^. Changes in sodium concentration may also be indirectly linked to hypoxia and oxidative phosphorylation as a result of both the relationship between sodium-hydrogen exchange and pH, and the energetic requirements for the transmembrane sodium-potassium pump^12,13^. The important role of pH in renal tumours is exemplified by the extracellular expression of carbonic anhydrase IX (CAIX) as a pH regulator^35^, and can be demonstrated using PET as well as on immunohistochemistry in the context of ccRCC^36^. Metabolic rewiring of renal cancer can be indirectly imaged using ^99^Tc-sestamibi with single-photon emission computed tomography (SPECT) which detects high mitochondrial density^37^. This is a characteristic of oncocytic neoplasms where defective mitochondria accumulate due to an impaired respiratory complex I, resulting in hypoxia^38^. However, these techniques involve ionising radiation and cannot differentiate between RO and chRCC with high certainty^39^. Therefore, an MRI-based approach to discriminate these offers a potential tool to address this clinical unmet need.

The limitations of this study include a small patient cohort size, justifiable in a proof-of-concept study. The low spatial resolution limited the ability to differentiate the medulla and cortex separately as previously observed, and therefore changes in the cortico-medullary gradient could not be assessed^18,19^. We have also not corrected for cystic/necrotic regions in tumours which may have affected the TSC quantification, but we did use the ^1^H-MRI derived biomarkers to corroborate the ^23^Na-MRI results.

In conclusion, by detecting differential sodium concentrations within RO and chRCC, and by correlating the TSC with *R*_2_*, we have shown the feasibility and potential of using ^23^Na-MRI to differentiate benign from malignant oncocytic renal tumours. Future larger studies should evaluate whether the technique could be implemented as a routine imaging tool after biopsy to noninvasively characterise the entire tumour and thus predict aggressiveness, which the biopsy cannot.

## Data Availability

All data produced in the present study are available upon reasonable request to the authors

## Supplementary

**Supplementary Table 1:**
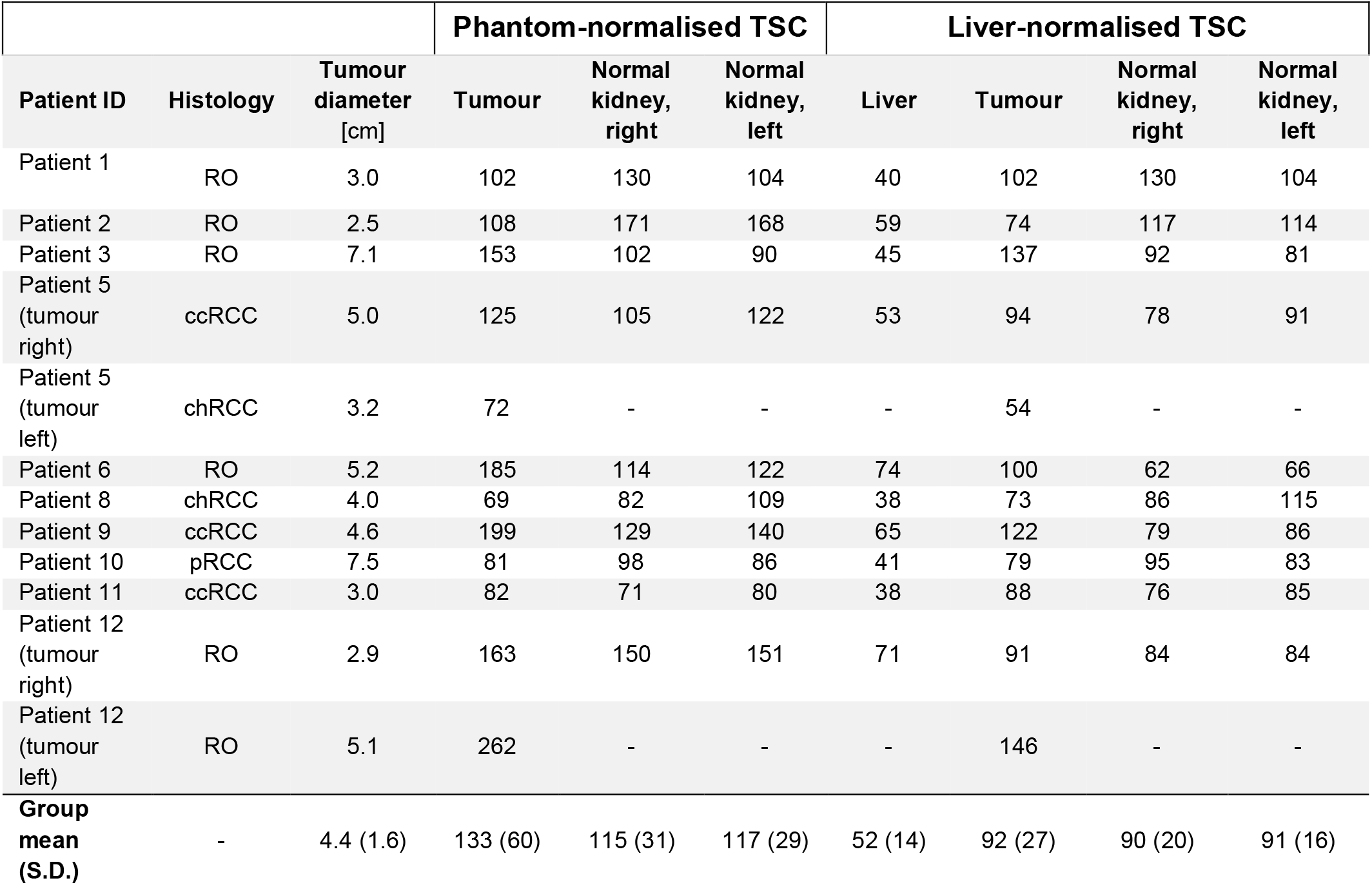
TSC quantification results of kidney tumours and bilateral normal kidney parenchyma from participating patients. RCC = renal cell carcinoma, TSC = total sodium concentration. Units of TSC are in mM.

**Supplementary Figure 1:**
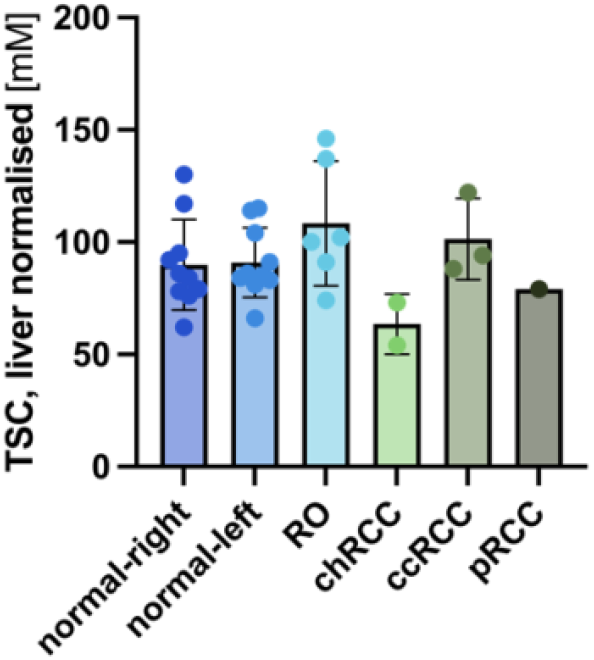
Barplot presenting the B_1_-corrected mean TSC from normal kidney parenchyma and across the kidney tumour subtypes, normalised to liver TSC signal.

**Supplementary Table 2:** Quantification of *R*_2_* derived from ^23^Na-MRI and ^1^H-MRI in tumours and normal kidney parenchyma. Units are s^-1^.

| Patient ID | <sup>23</sup>Na-MRI derived $R_2^*$ | | | <sup>1</sup>H-MRI derived $R_2^*$ | | |
| --- | --- | --- | --- | --- | --- | --- |
|  | Tumour | Normal kidney, right | Normal kidney, left | Tumour | Normal kidney, right | Normal kidney, left |
| Patient 3 | - | - | - | 16 | 20 | 20 |
| Patient 5 (tumour right) | - | - | - | 28 | 22 | 32 |
| Patient 5 (tumour left) | - | - | - | 30 | - | - |
| Patient 6 | 76 | 49 | 49 | 16 | 22 | 21 |
| Patient 8 | 64 | 41 | 48 | 14 | 21 | 17 |
| Patient 9 | 46 | 57 | 62 | 13 | 20 | 23 |
| Patient 10 | 79 | 40 | 46 | 48 | 18 | 19 |
| Patient 11 | 50 | 60 | 57 | 13 | 22 | 20 |
| Patient 12 (tumour right) | 54 | 48 | 48 | 14 | 17 | 17 |
| Patient 12 (tumour left) | 54 | - | - | 12 | - | - |
| **Group mean (S.D.)** | 60 (13) | 49 (8) | 51 (6) | 19 (13) | 20 (2) | 19 (2) |

**Supplementary Table 3:** Quantification of IVIM DWI derived parameters *f*_p_ (in arbitrary units) and *D*_0_ (in × 10^−3^ mm^2^ s^-1^) in tumours and normal kidney parenchyma.

| | $f_p$ | | | $D_0$ | | |
| --- | --- | --- | --- | --- | --- | --- |
| Patient ID | Tumour | Normal kidney<br>parenchyma,<br>right | Normal kidney<br>parenchyma,<br>left | Tumour | Normal kidney<br>parenchyma,<br>right | Normal kidney<br>parenchyma, left |
| Patient 6 | 0.034 | 0.075 | 0.051 | 2.264 | 2.106 | 2.144 |
| Patient 8 | 0.085 | 0.057 | 0.044 | 1.832 | 2.542 | 2.606 |
| Patient 9 | 0.097 | 0.006 | 0.019 | 2.071 | 1.826 | 1.933 |
| Patient 10 | 0.045 | 0.007 | 0.022 | 0.970 | 2.061 | 2.066 |
| Patient 11 | 0.154 | 0.066 | 0.059 | 2.279 | 2.066 | 2.028 |
| Patient 12<br>(tumour right) | 0.028 | 0.060 | 0.091 | 2.082 | 2.115 | 2.047 |
| Patient 12<br>(tumour left) | 0.028 | - | - | 2.348 | - | - |
| **Group mean**<br>**(S.D.)** | 0.067<br>(0.047) | 0.045 (0.031) | 0.048 (0.027) | 1.978<br>(0.477) | 2.120 (0.233) | 2.138 (0.239) |

